# Gene dysregulation among virally suppressed people living with HIV links to non-AIDS defining cancer pathways

**DOI:** 10.1101/2024.01.03.24300792

**Authors:** Bryan C. Quach, Eric Earley, Linran Zhou, Caryn Willis, Jesse A. Marks, Jeran K. Stratford, Fang Fang, Laura J. Bierut, M-J S. Milloy, Kanna Hayashi, Kora DeBeck, Dana B. Hancock, Ke Xu, Bradley E. Aouizerat, Eric O. Johnson

## Abstract

Combination antiretroviral therapy (ART) has changed the landscape of the HIV epidemic by providing an effective means for viral suppression to people living with HIV (PLWH). Understanding living with HIV as a chronic disease requires an improved understanding of how HIV and/or ART impacts susceptibility to and development of co-occurring conditions. Genome-wide gene expression (transcriptome) differences provide a key view into biological dysregulation associated with living with HIV. Here we present the first whole blood transcriptome-wide study comparing gene expression profiles between virally suppressed PLWH and HIV negative individuals (N=555). We identify 566 genes and 5 immune cell types with differential proportions by HIV status, which were significantly enriched for immune function and cancer pathways. Leveraging quantitative trait loci (QTL) for these HIV status-associated genes, partitioned heritability, and colocalization analyses, we observed limited genetic drivers of these relationships. Our findings suggest that gene dysregulation does not return to a pre-infection state for virally suppressed PLWH, and that persistent gene dysregulation is broadly associated with immune function and cancer pathways, highlighting potential biological drivers for these causes of excess mortality and targets for pharmacological preventative treatment among PLWH.

## Introduction

Advancements in HIV treatment have transformed infection with the retrovirus into a chronic disease in much of the Western World. For people living with HIV (PLWH), adherence to an effective combination antiretroviral therapy (ART) regimen suppresses viral replication and reduces the risk of progression to Acquired Immune Deficiency Syndrome (AIDS)^1^. However, with improved survival, PLWH on ART are experiencing increased rates of non-AIDS defining conditions (e.g., cardiovascular disease, diabetes mellitus, neurocognitive disorders)^2,3^ Among these diseases, non-AIDS defining cancers, such as Hodgkin’s lymphoma and lung, liver, kidney, and anal cancers, are key drivers of excess mortality for PLWH^4–6^. Persistent HIV-associated inflammation is hypothesized to contribute to the pathogenesis of non-AIDS defining conditions and chronic, virologically suppressed HIV disease or ART may accelerate cellular aging that influences morbidity and mortality risks for PLWH^7,8^.

Studies of host gene expression changes in HIV have uncovered a wealth of molecular targets associated with HIV acquisition, acute infection, progression, and latency.^9,10^ A handful of these studies investigated aberrant gene expression in B-cells or lung biopsy samples for both AIDS-defining and non-AIDS-defining cancers within PLWH^9,10,11^. No studies to date have measured gene expression differences transcriptome-wide in virally non-detectable PLWH compared to HIV negative individuals. The extent to which differential gene expression in PLWH is associated with HIV-associated cancers remains to be studied.

To investigate gene dysregulation associated with successfully treated HIV, we conducted the first transcriptome-wide association study (TWAS) comparing virally non-detectable PLWH (N=194) to HIV-1 seronegative individuals (N=361). By using RNA sequencing of whole blood instead of isolated immune cells or cell lines, this study captures a broad, cross-cell-type signature of gene expression changes *in vivo* for chronic HIV disease and ART: identifying dysregulated genes in PLWH linked to multiple aspects of immune function and cancer-related biological pathways. To examine cell-type specific effects, we applied gene expression deconvolution and tested for differential cell-type proportions by HIV status. Additionally, we assessed cell-type specific expression for HIV-associated genes from our TWAS using expression profiles available in the Database of Immune Cell Expression, Expression quantitative trait loci, and Epigenomics (DICE)^12^. We characterized genetic contributions to dysregulated genes by mapping expression quantitative trait loci (eQTL) and assessed genetic relationships between HIV-associated genes and immune-related traits and multiple cancers using partitioned heritability enrichment and colocalization analyses. This study provides insight into the interplay of chronic HIV infection, ART, and genetically-regulated gene expression as it relates to molecular processes and pathways linked to comorbid conditions, including non-AIDS defining cancers. The identified genes, and drugs targeting them, may be key to developing treatments for co-occurring health conditions in PLWH.

## Results

### Differential gene expression by HIV status

Gene expression was measured using paired-end RNA-seq from whole blood samples of 194 PLWH and 361 people seronegative for HIV-1 (Table 1). All PLWH met the Center for Disease Control and Prevention (CDC) definition of virally non-detectable (i.e., <200 viral copies/mL of blood)^13^ and were on ART at the time of blood draw. We conducted transcriptome-wide differential expression analysis by HIV status, accounting for potential biological and technical sources of confounding (see *Online Methods*). Of 20,211 genes tested, we identified 566 differentially expressed genes (DEGs; permutation p-value <0.05; Figures 1A and 1B, Supplemental Table 1). Relative to HIV-negative samples, 274 DEGs were up-regulated and 292 were down-regulated in PLWH, and we did not observe a significant bias towards a higher proportion of up-regulated or down-regulated genes (Binomial test *p*=0.48).

**Figure 1.**
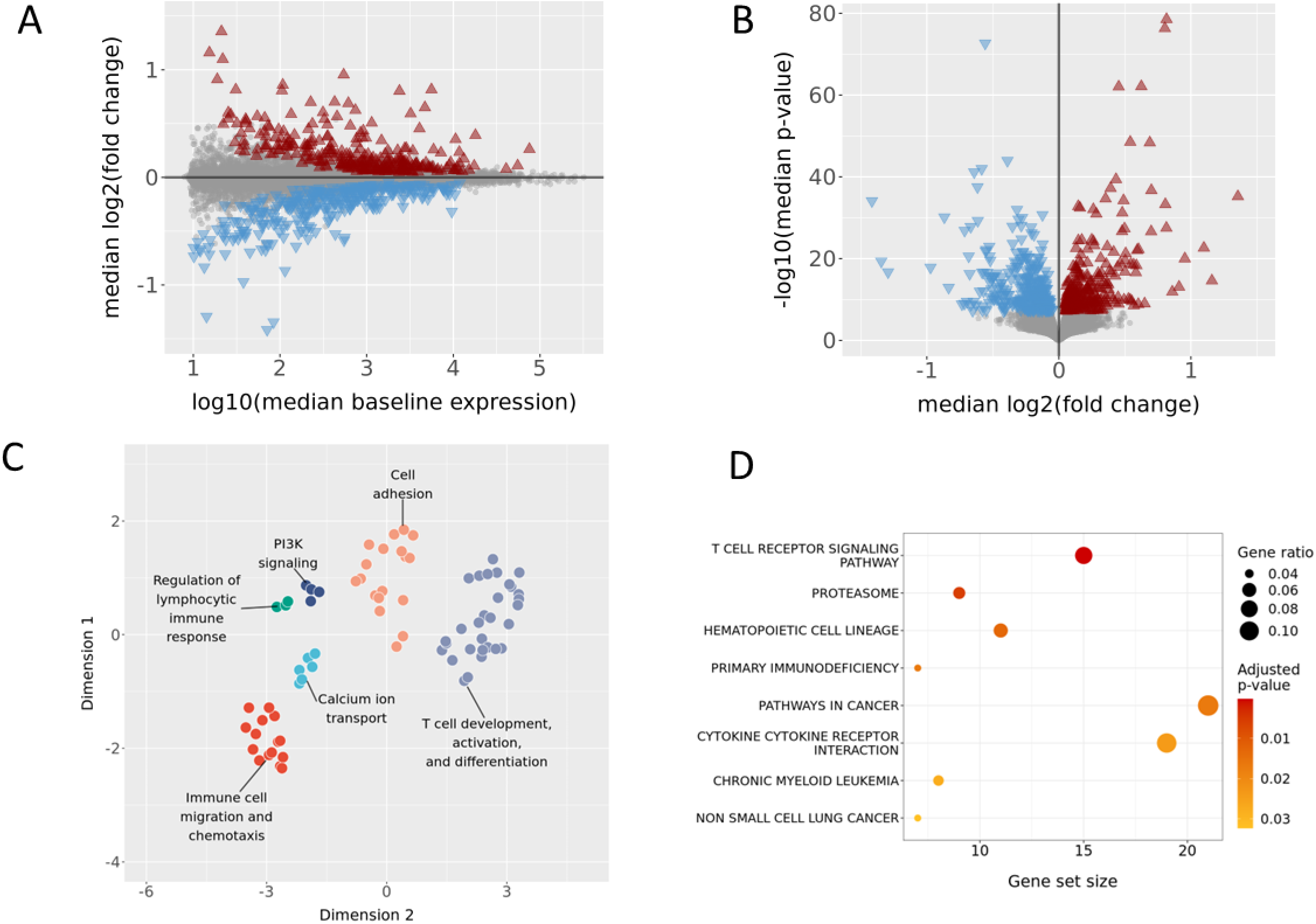
Differentially expressed genes by HIV status are overrepresented in immune processes and cancer pathways. A) The median of the average gene expression level and fold change values across 1000 bootstrap sampling iterations for each gene tested for differential expression by HIV status. Significantly up-regulated and down-regulated genes at FDR <0.05 are color-coded red and blue, respectively. Genes not passing the significance threshold are shown as gray points. B) The median of p-values and fold change values across 1000 bootstrap sampling iterations for each gene tested for differential expression by HIV status. Significantly up-regulated and down-regulated genes at FDR <0.05 are color-coded red and blue, respectively. Genes not passing the significance threshold are shown as gray points. C) UMAP projection of semantic distances between GO biological process terms with overrepresentation of HIV status DEGs. GO term clusters identified by HDBSCAN (distinguished by color) are manually assigned labels denoting an underlying theme for terms within a cluster. D) Dotplot of significant (FDR <0.05) KEGG canonical pathway terms from gene set overrepresentation analysis for all HIV status DEGs. The gene ratio indicates the proportion of DEGs in the gene set for a KEGG term relative to the gene set size.

**Table 1.** Sample characteristics and demographics of study participants.

|  |  | HIV- | HIV+ |
| --- | --- | --- | --- |
| **Total N** |  | 361 | 194 |
| **Sex** |  |  |  |
|  | *Male, N (%)* | 265 (73.4) | 148 (76.3) |
|  | *Female, N (%)* | 96 (26.6) | 46 (23.7) |
| **Mean age (SD)** |  | 48.96 (10.25) | 51.07 (8.37) |
| **STD status** |  |  |  |
|  | *Never, N (%)* | 90 (24.9) | 27 (13.9) |
|  | *Ever, N (%)* | 271 (75.1) | 167 (86.1) |
| **Mean RNA sample RIN (SD)** |  | 7.13 (0.52) | 7.07 (0.55) |
| **log10 viral copies/mL (SD)** |  | 1 (0) | 1.58 (0.12) |
| **Total (%)** |  | 361 (65.05) | 194 (34.95) |

Percentages and standard deviations (SD) for specific characteristics are in relation to the respective HIV status group for a given table cell.

Among the HIV-associated DEGs, gene set overrepresentation analysis using the GO Biological Process ontology identified 77 significantly enriched GO terms (FDR <0.05), which we grouped into categories based on semantic similarity (Figure 1C, Supplemental Table 2). The GO terms reflected a central theme of dysregulation in immune system processes related to cell migration, cell adhesion, T cell biology, lymphocyte immune response, calcium signaling, and phosphatidylinositol 3-kinase (PI3K) signaling. Overrepresentation analysis using the MSigDB KEGG canonical pathways collection identified eight significantly enriched pathways (FDR <0.05; Figure 1D, Supplemental Table 3), several of which are related to immune function, including *T cell receptor signaling pathway*, *proteasome*, *hematopoietic cell lineage*, *primary immunodeficiency*, and *cytokine-cytokine receptor interaction*. Furthermore, three cancer pathways exhibited overrepresentation of HIV-associated DEGs: *pathways in cancer*, *chronic myeloid leukemia*, and *non-small cell lung cancer*. We note that the DEGS in *chronic myeloid leukemia* and *non-small cell lung cancer* are subsets of the *pathways in cancer* gene set (Supplemental Figure 1). This gene set, which encompasses several biological pathways, is related to multiple types of cancer and tumorigenesis (Supplemental Figure 2)^14–23^.

### Cell-type proportion differences by HIV status and cell-type specific expression

Deconvolution of whole blood data was performed using the LM22 leukocyte reference panel^24^ to estimate the cell-type proportions of 22 cell types. Of these, we observed five cell types with significant differences in proportions by HIV status (Bonferroni *p* < 0.05; Figure 2A). The mean proportions of CD8+ T cells and follicular helper T cells were higher in HIV+ samples. In contrast, the mean proportions of naïve CD4+ T cells, resting CD4+ memory T cells, and resting mast cells were lower in HIV+ samples. We further characterized cell-type specific expression across 15 immune cell types for the HIV-associated DEGs. We examined differences in expression profiles using data from DICE, which characterizes immune cell gene expression in a healthy state^12^. For each cell type, the mean of transcripts per million (TPM) values across samples is used to represent the expression level for a gene. To assess which cell types exhibited similar/dissimilar expression profiles based on the overlap between HIV-associated DEGs with data available in DICE, we applied Pearson correlation hierarchical clustering of DICE expression data for the DEGs. The highest divergence in expression was between the T lymphocyte subset and innate immune cell types. Within the T lymphocytes, the cell types further grouped by naïve and memory T cells, with activated CD4+ and CD8+ T cells being an outgroup (Figure 2B). This clustering structure resembles the global unsupervised hierarchical clustering using the top 1000 most variable genes in DICE,^12^ suggesting that DEGs in the current study align with overall gene expression landscape in immune cells.

**Figure 2.**
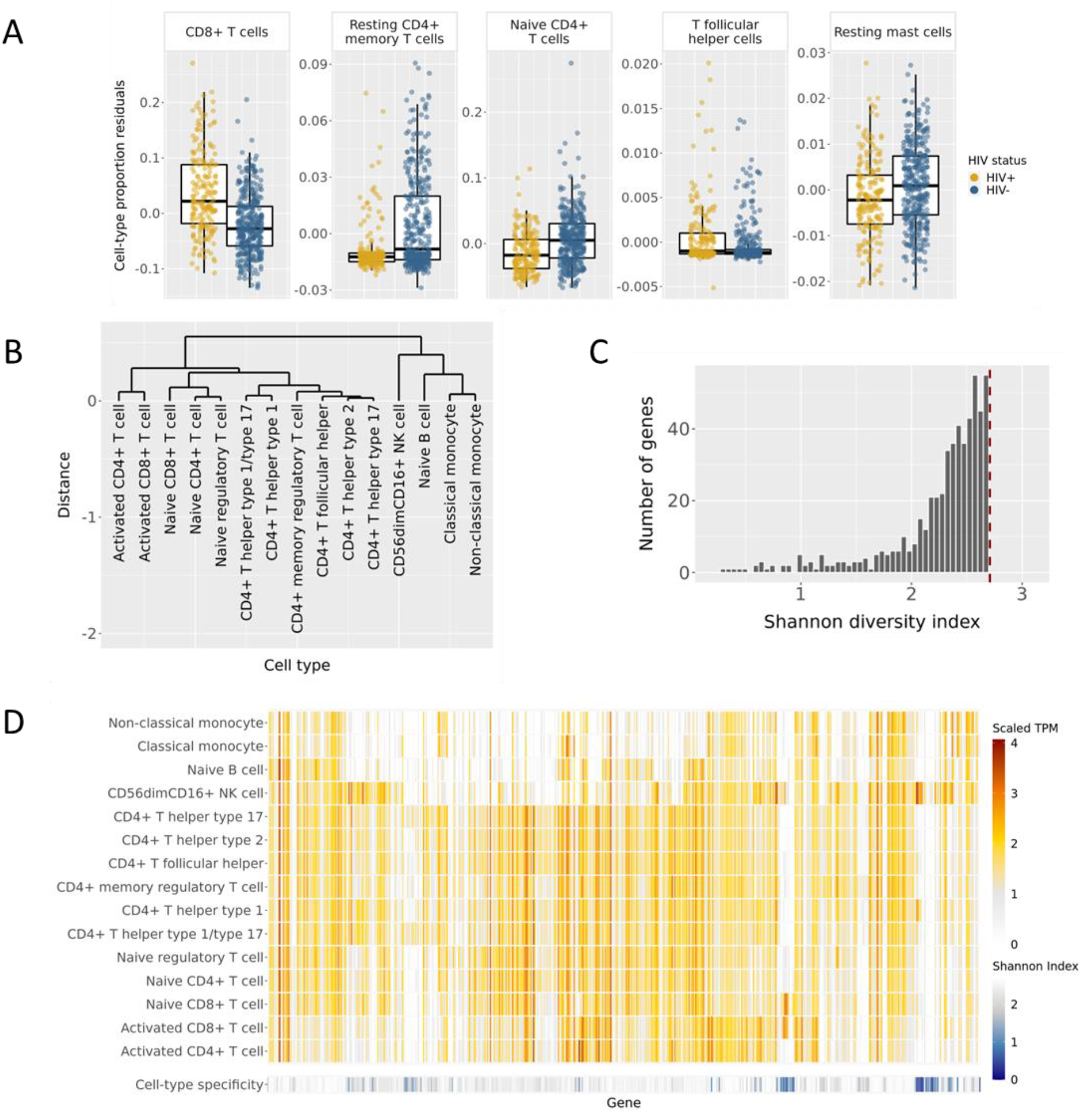
Gene dysregulation in PLWH impacts multiple immune cell types. A) Residuals for estimated proportions from the LM22 cell-type reference panel after removing potential confounding effects of biological variables (see Methods). Only cell types with a significant difference by HIV status (Bonferroni adjusted p <0.05) are shown. See Supplemental Figure X for all LM22 cell types. B) Hierarchical clustering of DICE cell types based on Pearson correlations between cell types using expression levels for the HIV-associated DEGs. C)Distribution of Shannon diversity index values for HIV-associated DEGs. The Shannon index values measure uniformity of gene expression levels across 15 immune cell types available from the DICE data repository. Higher values indicate more uniform expression levels across cell types. The red dashed horizontal line indicates the theoretical maximum value for these data (i.e., uniform gene expression levels across all cell types). D) Gene expression levels in 15 DICE immune cell types for HIV status DEGs. Gene expression levels are displayed as log10(mean TPM +1) values. Shannon diversity index values are displayed for each gene to denote the degree of cell-type specificity (larger values equate to more uniform expression across cell types).

To assess the degree of cell-type specificity for each HIV-associated DEG, we calculated Shannon diversity index values^25^ based on the proportion of total expression (sum of cell type mean TPMs) for each cell type. The distribution of Shannon index values was strongly left-skewed, indicating that most DEGs were expressed more uniformly across the 15 cell types (Figure 2C and 2D). When examining genes in the fifth percentile for Shannon index values (i.e., less evenness), most of these DEGs had high mean TPM proportions concentrated within only one or two cell types (Supplemental Figure 3A). For example, the antigen encoded by *KLRD1* is preferentially expressed on natural killer (NK) cells^26^, and we observed that *KLRD1* was disproportionately expressed in CD56^dim^ CD16^+^ natural killer (NK) cells with low expression levels in other cell types. However, in addition to the disproportionately high expression cell types, some genes still had moderate or high expression levels across several other cell types (Supplemental Figure 3B). *CD40LG*, a gene well-studied in HIV infection and immunity^27^, exhibited disproportionately strong expression in naïve activated CD4+ T cells but also showed moderate expression in other CD4+ T cell subtypes and activated CD8+ T cells. These findings show that gene dysregulation in virally suppressed PLWH occurs in multiple cell types involved in both innate and adaptive immune response.

### Characterization of gene dysregulation by ART status

Since effective ART suppresses viral replication and stalls HIV disease progression, we sought to better understand whether our HIV-associated DEGs were impacted by ART exposure. We used work by Massanella *et al.*^28^ to define HIV-associated DEGs as ART-affected or ART-unaffected. Massanella *et al.* reported 4,157 genes with differential expression in peripheral blood mononuclear cell samples from 32 males living with HIV pre-ART and 48 weeks post-ART initiation. Intersecting our HIV-associated DEGs with this gene list resulted in 234 HIV-associated DEGs being classified as ART-affected and 332 DEGs classified as ART-unaffected (Supplemental Table 1). Among ART-affected HIV-associated DEGs, a higher proportion were down-regulated (Binomial test *p*=0.011). No significant enrichment in proportion of up- or down-regulated genes was observed for ART-unaffected, HIV-associated DEGs (Binomial test *p*=0.25; Figure 3A). Comparing fold change values reported by Massanella *et al.* (post-ART treatment vs. pre-ART treatment) to the fold changes for the ART-affected, HIV-associated DEGs (HIV+ vs. HIV-) revealed that almost all DEGs have an inverse direction of expression change between studies (Figure 3B). 137 genes that were upregulated post-ART compared to pre-ART still exhibited lower expression for the HIV+ group relative to HIV-samples. Similarly, the 97 post-ART downregulated DEGs showed higher expression for the HIV+ group relative to the HIV-samples. This discordance in direction of effect suggests that ART and viral suppression shifts expression of most DEGs toward the HIV uninfected state but does not fully restore gene regulation to this state.

**Figure 3.**
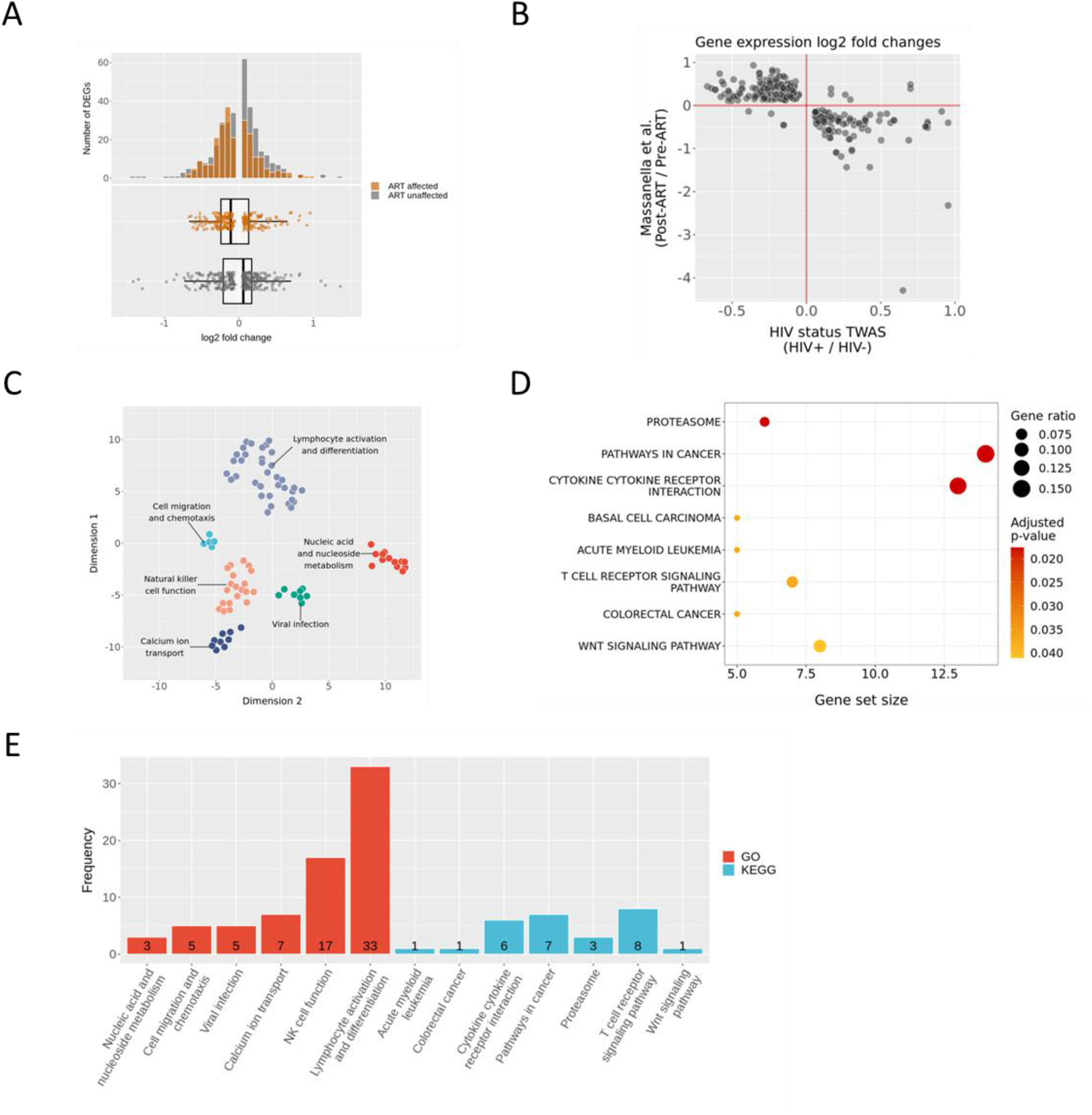
HIV-associated genes are affected by ART. A) Distribution of log2 fold change values for HIV-associated DEGs partitioned by ART status. B) Comparison of differential expression analysis log2 fold changes between this study and Massanella et al. for ART-affected, HIV-associated DEGs. The log2 fold change values plotted from this study are the medians of the log2 fold change values for each gene across 1000 bootstrapping iterations using 80% of the sample size while preserving the HIV+ to HIV-ratio. C) UMAP projection of semantic distances between GO biological process terms with overrepresentation of ART-affected, HIV-associated DEGs. GO term clusters identified by HDBSCAN (distinguished by color) are manually assigned labels denoting an underlying theme for terms within a cluster. D) Dotplot of significant (FDR <0.05) KEGG canonical pathway terms from gene set overrepresentation analysis for ART-affected, HIV-associated DEGs. The gene ratio indicates the proportion of DEGs in the gene set for a KEGG term relative to the gene set size. E) Number of ART-unaffected, HIV-associated DEGs belonging to GO biological process and KEGG canonical pathway terms from gene set overrepresentation analysis for ART-affected, HIV-associated DEGs.

ART-affected, HIV-associated DEGs were overrepresented for 90 GO biological process terms (FDR <0.05; Figure 3C) and 8 MSigDB KEGG canonical pathway terms (FDR <0.05; Figures 3D and 3E). In line with the full set of HIV-associated DEGs, many of the significant GO terms were related to immune function processes, but some processes were distinctly enriched for these ART-affected DEGs such as nucleic acid and nucleoside metabolism, a critical component of reverse-transcriptase inhibition through ART. Other processes targeted by ART were also overrepresented, including terms related to viral infection (e.g., *regulation of viral life cycle* and *viral process*). Many biological processes were also related to NK cell functions such as cell killing, ruffling, inositol lipid-mediated signaling, and chemokine signaling^29,30^. Several KEGG pathway terms in the overrepresentation analysis for all DEGs were also significant when assessing only ART-affected DEGs, with *pathways in cancer* persisting as the term enriched for the most DEGs (Supplemental Table 4). Of the signaling pathways in the *pathways in cancer* gene set, all the pathways described above, including Wnt/β-catenin, PI3K/AKT, MAPK/ERK, p53, and TGF-β signaling, had at least one ART-affected HIV-associated DEG (Supplemental Figure 2). ART-unaffected, HIV-associated DEGs were overrepresented for 5 GO biological process and 1 KEGG canonical pathway term (FDR <0.05; Supplemental Table 5) more narrowly linked to T cell biology (e.g., *T cell receptor signaling pathway, T cell selection*, *T cell co-stimulation*, *cell-substrate adhesion*). However, these findings do not preclude the involvement of ART-unaffected DEGs in the biological processes and pathways with an overrepresentation of ART-affected DEGs. Across the GO and KEGG term gene sets with overrepresentation of ART-affected DEGs, 80 ART-unaffected DEGs are components of 77 of the terms. For the GO biological process term clusters (Figure 3C), each cluster includes at least 1 term with an ART-unaffected DEG. For the KEGG terms, *basal cell carcinoma* is the only term without an ART-unaffected DEG. (Supplemental Table 6; Figure 3E). These results show that although ART influences a broad group of immune and cancer-related molecular processes, gene dysregulation in these processes are not solely attributable to ART.

### eQTL mapping for HIV status DEGs, partitioned heritability, and colocalization

Given the observed immune- and cancer-related biological processes and pathways, we assessed whether heritability of immune- and cancer-related traits was enriched in genomic regions for HIV-associated DEGs. We tested for partitioned heritability enrichment of 30 GWAS traits (see Supplemental Table 7 for GWAS details) using stratified LDSC (sLDSC) and the set of HIV-associated DEGs with a window of 100 kilobases (kb) upstream and downstream around each DEG. Of these traits, eight showed statistically significant heritability enrichment (Bonferroni *p* < 0.05; Figure 4A; Supplemental Table 8, Supplemental Figure 4). Three of the significant traits were blood cell counts (platelet, red blood cell, and white blood cell count). The other significant traits include asthma (estimated proportion of *h*^2^ [prop. *h*^2^]=0.1197), Crohn’s Disease (prop. *h*^2^=0.1069), rheumatoid arthritis (prop. *h*^2^=0.1731), primary biliary cirrhosis (prop. *h*^2^=0.1259), and primary sclerosing cholangitis (prop. *h*^2^=0.1594). These five traits are inflammatory diseases that impact several different tissues. Although 12 cancer GWAS traits were tested, none showed significant heritability enrichment for DEG regions. Likewise, enrichment was not observed for HIV acquisition, suggesting that collectively our observed HIV-associated DEGs are not strongly associated with known genetic risk for cancer or HIV acquisition.

**Figure 4.**
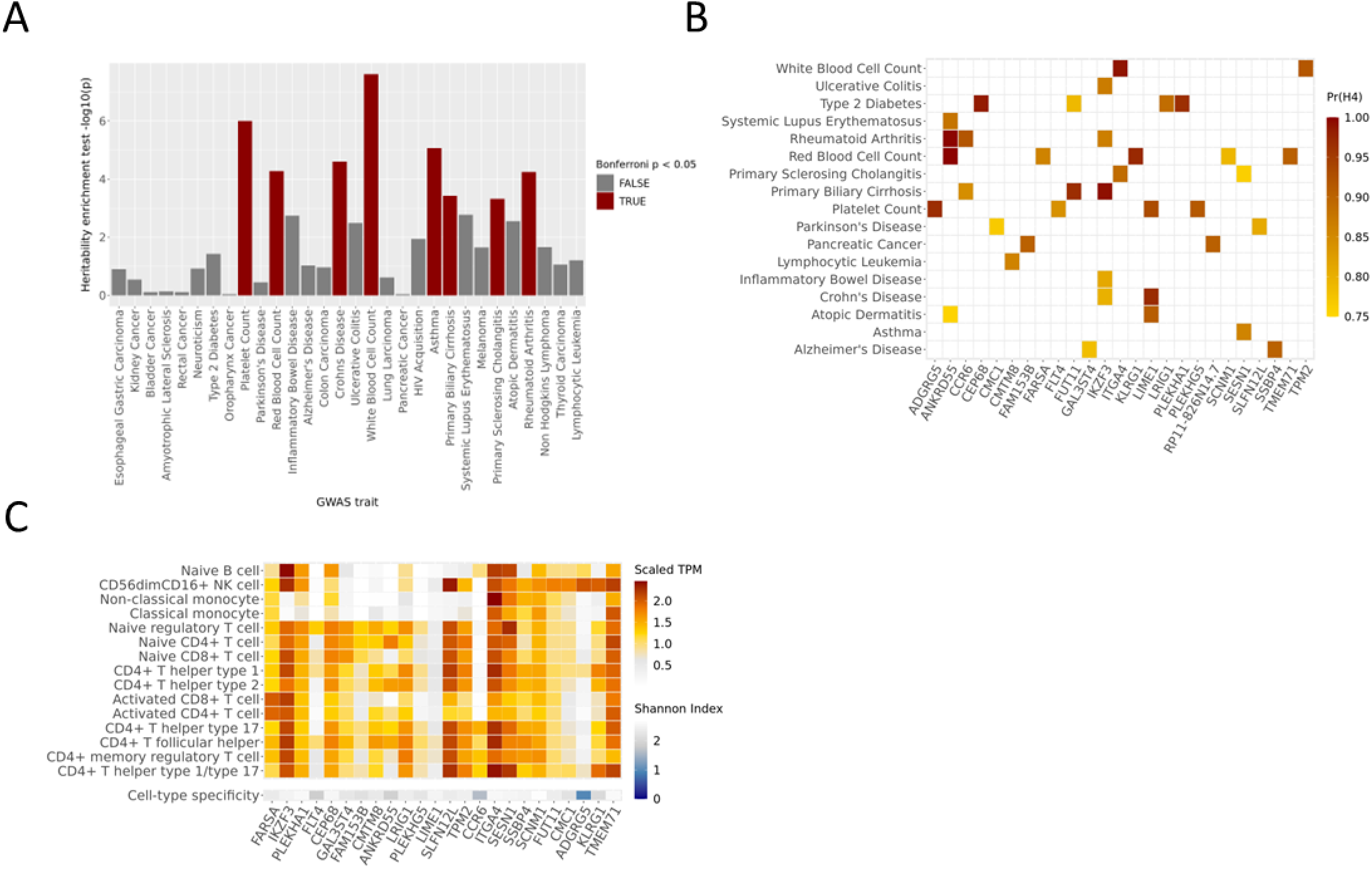
HIV-associated DEGs are not strongly associated with known genetic risk for cancer or HIV acquisition. A) Partitioned heritability enrichment test –log10 transformed p-values using sLDSC for 30 GWAS traits. B) Posterior probabilities for genes (x-axis) that colocalize with at least one of thirty immune-related and cancer GWAS traits. Only gene-trait pairs with a colocalization posterior probability >0.75 are shown here. The posterior probabilities correspond to the colocalization hypothesis that both a gene and trait are associated and share a causal variant. C) Gene expression levels in 15 DICE immune cell types for HIV-associated DEGs in Figure 4B. Gene expression levels are displayed as log10(mean TPM +1) values. Shannon diversity index values are displayed for each gene to denote the degree of cell-type specificity (larger values equate to more uniform expression across cell types). Gene *RP11-826N14.7* is not included due to not being available in DICE.

The statistically nonsignificant global enrichment results do not preclude some of the DEGs from having shared genetic signal with these traits. We explored this possibility by first conducting cis-expression quantitative trait locus (eQTL) mapping for the 566 HIV status DEGs using gene expression and imputed genotype dosage data from our study cohort. After applying a two-stage multiple testing correction (see *Online Methods*), we identified 22,475 eQTL corresponding to 258 eGenes (DEGs with at least one significantly associated genetic variant; Supplemental Table 9). Of these eGenes, 110 were ART-affected, 148 ART-unaffected, and five overlapped with the KEGG p*athways in cancer* gene set. Sixty-seven eSNPs (genetic variants with at least one significantly associated eGene) corresponding to four eGenes were *pathways in cancer* genes and designated as ART-affected. Fifty-three eSNPs were linked to a single ART-unaffected, *pathways in cancer* eGene, *LAMC3*. These *pathways in cancer* eGenes are components of Wnt/β-catenin, PI3K/AKT, MAPK/ERK, and TGF-β signaling (Supplemental Figure 2). These eSNP findings suggest genetic association with cancer risk among a subset of eGenes for both ART-affected and unaffected gene sets.

For the 258 eGenes, we used an 800 kb total *cis* window around the gene body to define regions to test for colocalization with each of the 30 GWAS traits from the partitioned heritability analysis (see Supplemental Table 10 for colocalization test summary statistics). As shown in Figure 4B, 17 traits and 26 eGenes had at least one colocalization event (posterior probability >0.75 of the trait and gene being associated and sharing a causal variant). Despite not observing heritability enrichment for cancer traits, we identified colocalization between genetic signals for pancreatic cancer and *FAM135B* and *RP11-826N14.7*, and lymphocytic leukemia and *CMTM8*. No colocalization tests between eGenes and HIV acquisition met our significance threshold. Additionally, we observed pleiotropic effects for several eGenes including *ANKRD55* (systemic lupus erythematosus, rheumatoid arthritis, red blood cell count, and atopic dermatitis), *CCR6* (rheumatoid arthritis and primary biliary cirrhosis), *FUT11* (type 2 diabetes and primary biliary cirrhosis), *IKZF3* (ulcerative colitis, rheumatoid arthritis, primary biliary cirrhosis, inflammatory bowel disease, and Crohn’s disease), *ITGA4* (white blood cell count and primary sclerosing cholangitis), *LIME1* (platelet count, atopic dermatitis, and Crohn’s disease), and *SESN1* (primary sclerosing cholangitis and asthma). Based on cell-type gene expressions in DICE, these pleiotropic eGenes showed varying levels of cell-type specificity (Figure 4C). For example, *ITGA4*, and *SESN1* showed ubiquitous expression, whereas *IKZF3* displayed expression in lymphocyte cell types only, which aligns with its well-established role as a regulator of lymphocyte differentiation^31^. *ANKRD55* exhibited CD4+ T cell specific expression (noted previously in Lopez de Lapuente *et al.*^32^), and *CCR6* showed expression in only naïve B cells and a subset of CD4+ T cell types. Together our partitioned heritability and colocalization analysis results show that the genetic architecture of HIV-associated DEGs primarily overlaps with inflammatory diseases, but shared genetics between these DEGs and the cancers we assessed is very limited or not observed.

### Drug target analysis

To prioritize those DEGs that may have a higher potential for clinical translation, we sought to understand whether the HIV-associated DEGs were known targets for drug compounds in the clinical trials or approved drug development stage. From querying these DEGs against four drug target databases (DrugBank [go.drugbank.com], Open Targets [www.opentargets.org], Pharos [pharos.nih.gov], and Therapeutic Target Database [TTD; db.idrblab.net/ttd]), we observed 111 DEGs that were targets for at least one compound. Of these 111 DEGs, 55 were part of our “ART-affected” classification, and 42 were drug targets for an approved drug. Eleven of the *pathways in cancer* DEGs were drug targets. Three of the 11 were also eGenes, which highlights their potential for genotype-drug interactions (Figure 5; Supplemental Table 11).

**Figure 5.**
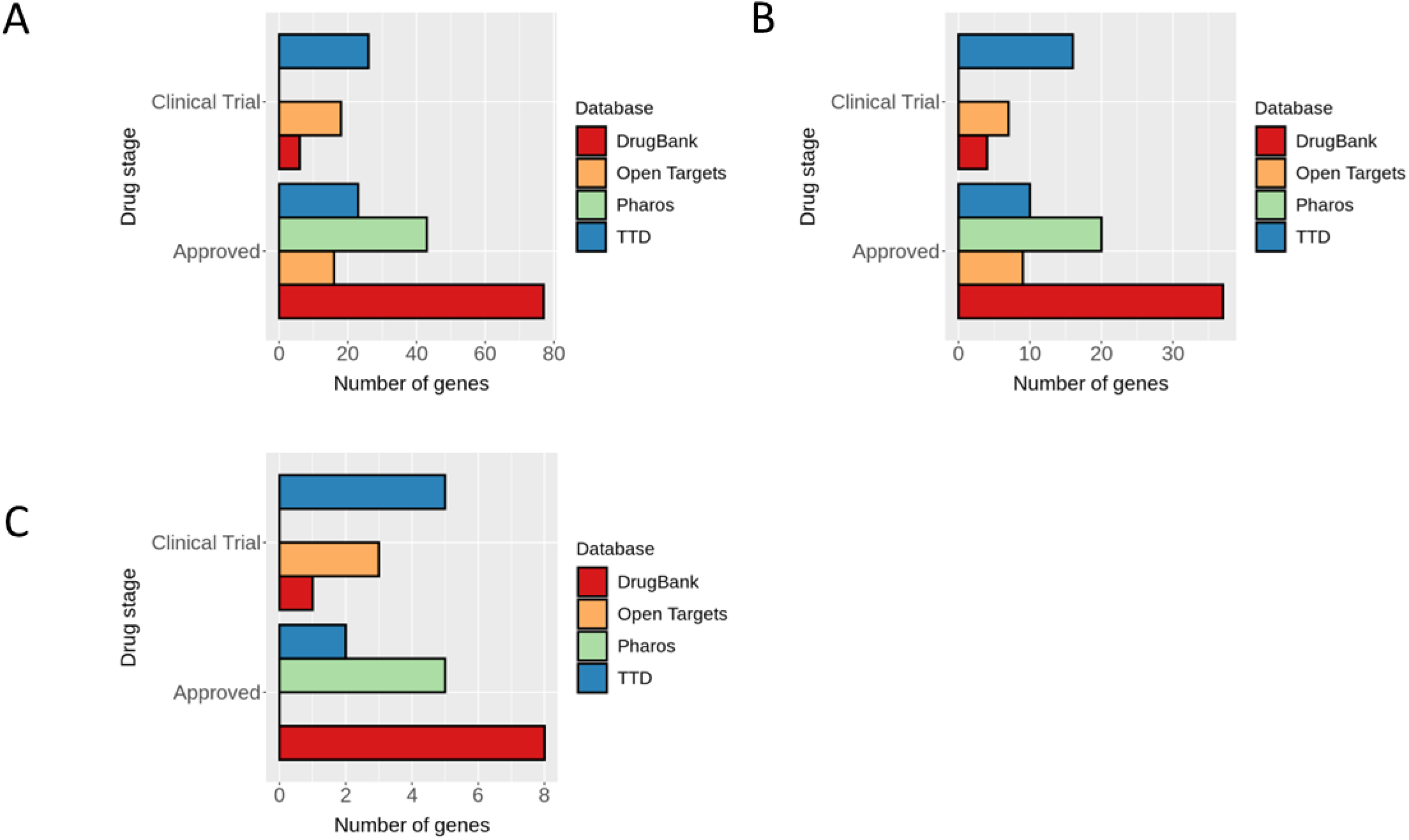
HIV-associated DEGs are targets for FDA-approved drugs or compounds in clinical trials. Most advanced drug development stage in drug target databases for drug-targeted, HIV status DEGs. A DEG may be present across multiple databases. Consequently, each bar does not represent an independent set of genes. A) All DEGs that are targets for at least one drug (N=111). B) ART-affected DEGs that are targets for at least one drug (N=55). C) KEGG *pathways in cancer* DEGs that are targets for at least one drug (N=11).

Through this analysis, we noted multiple types of potential interaction effects. For example, *PRKCA*, one of the ART-affected eGenes and HIV-associated DEGs involved in non-canonical *Wnt* signaling and MAPK/ERK signaling pathways, is targeted by midostaurin, a drug approved for treatment of acute myeloid leukemia and systemic mastocytosis. *PRKCA* is also targeted by bryostatin 1, which has been in clinical trials for Alzheimer’s disease and anti-cancer treatment. Bryostatins have also been observed to mildly inhibit acute HIV infection *in vitro* and activate latent HIV in monocytes and lymphocytes^33,34^.

## Discussion

Prior research to understand transcriptomic dynamics within the context of HIV infection has focused on changes during acute infection. In contrast, this study focused on our increasing need to understand HIV disease as a chronic condition, being the first to evaluate transcriptional alterations genome-wide among PLWH that are virally suppressed on ART. This large TWAS in whole blood identified 566 DEGs which show significant enrichment for immune system and cancer pathways – a substantial contributor to excess mortality among PLWH. Identification of eQTLs for DEGs and their assessment in partitioned heritability and colocalization analyses indicated that the observed pathway enrichments are generally not driven by underlying genetic susceptibility, suggesting the state of gene dysregulation related to these outcomes may be malleable with treatment. Intersecting this study’s 566 DEGs with results from the singular pre/post study of the gene expression effects of ART among men LWH, we found that approximately 40% of the HIV-associated DEGs may be affected by ART. Evaluation of drug target databases identified many DEGs that are targets of one or more FDA approved drugs and investigational compounds. Overall, these results suggest that gene dysregulation among virally suppressed PLWH may be a key focus for understanding the biological linkage between HIV and excess risk of non-AIDS defining cancers, as well as suggesting studies of potential targets for pharmacological interventions.

Although previous HIV transcriptomics studies exist, they involve only tens of samples which is substantially smaller than our study (N=555). Moreover, there are substantial variations in experimental designs (cell culture models, acute infection studies, differing tissue or cell type of interest, differences in gene expression assay technology, etc.)^35^ and statistical approaches which limits the feasibility of using prior studies to replicate the DEGs we identified or to evaluate prior studies finding in our results. However, we applied a robust approach to identifying DEGs by leveraging data-driven estimation of confounding variables and applying bootstrapping for our TWAS with a conservative multiple testing correction threshold, which supports confidence in our results.

Our observations of differential gene expression in chronic HIV disease relate to multiple elements of immune function. This aligns with clinical observations of persistent inflammation and immune dysfunction in PLWH that can affect treatment options and plans of care for co-occurring conditions^3,36^. GO terms and biological pathways with an overrepresentation of HIV-associated DEGs highlight multiple facets of immunity. Chemotaxis and cell migration processes are vital in directing leukocyte localization^37^. Calcium ion transport and signaling is involved in numerous immune processes such as lymphocyte development^38^, T cell and B cell receptor function^39,40^, mast cell activation^41^, and macrophage and dendritic cell phagocytosis^42^. Similarly, cell adhesion is critical to many immune processes including leukocyte homing to sites of inflammation, lymphocyte activation, and pathogen recognition and phagocytosis^43–46^. These clear connections between identified DEGs to expected dysregulation of the immune system under HIV supports confidence in our results.

### Dysregulated Signaling Pathways

Our findings also link HIV-associated gene expression to several molecular signaling pathways associated with cancer, e.g., Wnt/β-catenin, PI3K/AKT, MAPK/ERK, p53, and TGF-β signaling. Wnt/β-catenin signaling affects cell proliferation and is initiated by extracellular signals and Wnt proteins. Interactions between Wnt and cell membrane proteins regulate β-catenin, which can translocate from the cytoplasm to the nucleus to initiate transcription of target genes^47^. HIV-associated DEGs in the Wnt/β-catenin signaling pathway included *WNT7A*, *WNT10A*, *AXIN2*, *TCF7*, *LEF1*, *PPARD*, and *PRKCA*. *WNT7A* and *WNT10A* produce Wnt proteins; *AXIN2* contributes to a protein complex that degrades β-catenin in the cytoplasm; and *TCF7* and *LEF1* encode transcription factors TCF-1 and LEF-1, respectively, which recruit β-catenin to initiate gene transcription of target genes, including *PPARD*. *PRKCA* encodes protein kinase C alpha, which is involved in non-canonical Wnt signaling that can activate cell proliferation independent of β-catenin^48^. These genes all show lower mean expression among HIV+ samples, which largely indicates a repression of Wnt/β-catenin signaling pathway and cell proliferation. However, decreased expression of *AXIN2* implies decreased β-catenin degradation, allowing β-catenin to move into the nucleus. Knockdown experiments for the TCF-1 and LEF-1 transcription factors have been linked to disrupted T cell development and lymphoma in mice^49,50^.

PI3K/AKT signaling is a pathway related to cell proliferation and apoptosis. PI3K is an enzyme that phosphorylates the cell membrane phospholipid PIP2 to form PIP3 which activates the protein kinase AKT. AKT then interacts with many downstream targets to inhibit or activate various cellular processes. HIV-associated DEGs in this pathway included *LAMC3*, *ITGA6*, *PIK3R1*, *PIK3CB*, *PML*, *FOXO1*, and *CDKN1B*. *LAMC3* and *ITGA6* encode an extracellular matrix laminin subunit and plasma membrane integrin protein subunit alpha 6, respectively. Integrins function as receptors for laminins, and *ITGA6* activates PI3K with overexpression linked to multiple cancers^51,52^. *PIK3R1* and *PIK3CB* are genes for PI3K regulatory and catalytic subunits, respectively. *PML* encodes promyelocytic leukemia proteins that coordinate with the phosphatase PTEN to inhibit AKT^53^. *FOXO1* and *CDKN1B* are genes for targets of AKT, and their inhibition has been linked to tumorigenesis. AKT inactivates the FOXO1 transcription factor by phosphorylation, subsequently repressing apoptosis^54^. Similarly, AKT inactivates the cyclin-dependent kinase inhibitor p27 (i.e., the *CDKN1B* gene product), which increases cell proliferation^55^. Except for *PML*, the DEGs associated with PI3K/AKT signaling show decreased mean expression in HIV+ samples relative to HIV-samples, suggesting decreased PI3K activity. However, the decreased expression of FOXO1 and p27 genes implies AKT is still being activated, which may be occurring through PI3K-independent processes^56^.

In MAPK/ERK signaling, an extracellular signal leads to phosphorylation of the small GTPase Ras, resulting in a protein kinase cascade that activates ERK. Activated ERK interacts with transcription factors in the nucleus to promote cell proliferation. Ras can also promote cell proliferation by directly activating PI3K. HIV-associated DEGs involved in this pathway included *TGFA* (downregulated in HIV+ samples), *PRKCA* (downregulated), *RASSF1* (upregulated), and *DAPK3* (upregulated). *TGFA* encodes TGF-α, which binds to the plasma membrane epidermal growth factor receptor (EGFR) to active Ras. Protein kinase C alpha (product of *PRKCA*) phosphorylates Raf-1, a kinase downstream of Ras in the kinase cascade^57^. *RASSF1* produces multiple RASSF1 protein isoforms that can interact with Ras and mediate apoptosis^58,59^. *DAPK3* belongs to a gene family associated with apoptosis, and knockdown of DAPK3 has been shown to inhibit phosphorylation of ERK. Interestingly, DAPK3 exhibits dual properties as a tumor suppressor in some cell types and a cancer cell proliferator in others^60^.

The p53 signaling pathway is another pathway that regulates cell proliferation and apoptosis^61^. HIV-associated DEGs involved in this pathway included kinase inhibitor genes *CDKN2A* (upregulated) and *CDKN1B* (downregulated), protein kinase gene *CDK6* (upregulated), and *PML* (upregulated). *PML* was described above as an inhibitor of AKT in PI3K/AKT signaling, but in the context of p53 signaling, it functions both upstream and downstream of p53 tumor suppressor activation in the pathway. *PML* recruits p53 to nuclear bodies to enhance transcription of downstream target genes, but also is directly regulated by p53^62^. *CDKN2A* produces multiple protein isoforms, including tumor suppressor protein p14^ARF^ and kinase inhibitor p16. p14^ARF^ binds to ubiquitin-protein ligase MDM2 which prevents p53 from being degraded^63^. The p16 isoform inhibits the cell cycle progression protein complex CDK4/6 (for which *CDK6* encodes the catalytic subunit) from phosphorylating Rb. Phosphorylated Rb activates the E2F transcription factor, leading to cell proliferation. Similarly, p27 protein encoded by *CDKN1B* inhibits CDK2, another kinase that phosphorylates Rb.

In the TGF-β signaling pathway, only *TGFBR2* was identified as an HIV-associated DEG, downregulated in HIV+ samples. *TGFBR2* encodes one component of TGFBR, the transmembrane receptor protein heterodimer for TGF-β growth factor. When bound with TGF-β, TGFBR phosphorylates SMAD proteins that initiate a signaling cascade promoting cell growth. Notably, Liu et al. demonstrated that depletion of TGFBR2 stopped cancer progression in CD4+ T cells^64^.

### DEGs and cancer pathways

Incidence of non-AIDS defining cancers is higher among PLWH^4,65^ which aligns with our observations of HIV-associated DEGs being overrepresented in pathways that impact cell growth, proliferation, and apoptosis function. These processes are associated with many cancer types^15,16,18,66–68^. However, the direction of effect for the DEGs in these cancer-related pathways do not clearly confer a consistently increased or decreased cancer risk. For example, the observed decreased expression of *TCF7* and *LEF1* in PLWH in Wnt/β-catenin signaling would be expected to increase aberrant cell proliferation and cancer risk^49,50^. However, within the same pathway, the decreased expression of *WNT7A* and *WNT10A* would be expected to decrease cell proliferation, resulting in reduced cancer risk. These seemingly contradicting findings raise two important points. First, the directions of effect of our HIV-associated DEGs and their magnitude are based on mean group differences, so the behavior of these genes and related pathways in an individual may deviate from expectations based on the group means. Second, our findings are based on whole blood RNA-seq data and these DEGs may function differently in other tissues that may be more relevant for a given cancer type. Characterizing the impact of these genes in a specific context requires additional functional studies with refined hypotheses.

### Intersection of HIV and ART associated DEGs

Studying transcriptomic differences in PLWH on ART compared to HIV negative controls advances knowledge of persistent biological effects of living with HIV as a chronic condition. However, the confounding of chronic HIV infection and ART poses limitations in interpreting the impact of infection and treatment, separately. Clarifying the transcriptomic impact of antiretroviral drugs versus chronic HIV infection *in vivo* would benefit from peripheral blood RNA-seq studies in individuals using pre-exposure prophylaxis to prevent HIV infection. No publicly available data are available with this experimental design, so we used genes identified by Massanella *et al*.^28^ (comparing pre/post ART among PLWH) to categorize HIV-associated DEGs as ART-affected or unaffected. This comes with the caveat that Massanella *et al*. only included males with likely varying combinations of drug classes used for ART among participants. Nevertheless, we observed substantial overlap (40%) between DEGs associated with virally suppress PLWH in our study and DEGs associated with post/pre ART in Massanella *et al*.^28^ The predominant opposite direction of effect for these overlapping genes (e.g., upregulation in virally suppress PLWH compared to HIV-individual, compared with down regulation in post-vs. pre-ART among men LWH) suggests the expected drive of gene regulation toward the uninfected state under ART. Although ART-affected genes showed statistically significant enrichment for a number of pathways, including cancer-related pathways, and the ART-unaffected genes did not, a number of ART-unaffected genes also contribute to these significant pathways. Thus, while it appears that DEGs associated with ART influence a multitude of immune and cancer-related molecular processes, gene dysregulation in these processes cannot be exclusively attributable to ART. Follow-up functional studies will be needed to determine the mechanistic attribution of these observed associations.

### Cell-type deconvolution

Analysis of our HIV-associated DEGs using gene expression deconvolution reinforces several previously observed changes to leukocytes after HIV infection and ART treatment. An increase in CD8+ T cells and a decrease in CD4+ T cells has been well-documented^69–71^. However, prior literature provides a more complex picture for peripheral Tfh (pTfh) cells. Our observed increase in pTfh cells among PLWH has only been recorded in specific subtypes of pTfh cells (e.g., central memory CD27+CD45RO+ pTfh cells)^72,73^. Due to limitations of the cell type reference panel used for deconvolution of our whole blood gene expression data, estimating pTfh subtype proportions was not possible. The pTfh cell type is a major source of HIV latent reservoir that is highly susceptible to HIV infection^74^, so increases in pTfh cell proportion in virally suppressed PLWH on ART may indicate a larger HIV latent reservoir^75^. In relation to our finding of mast cell proportion differences by HIV status, we note that mature mast cells are tissue-resident, so our estimates from deconvolution may be relying on cell-type gene expression markers that lack the specificity to differentiate between mast cells and basophils or their common progenitors.

Regardless, a change in these cell populations warrants further experimental validation given that mast cells have been noted to harbor HIV reservoirs and basophils are involved in viral transmission to CD4+ T cells^76^. From analyzing the DICE single cell expression data for the HIV-associated DEGs, we noted genes expressed across the spectrum of cell type selectivity (e.g., *KLRD1* expressed predominantly in NK cells versus *CD40LG* expressed ubiquitously across T cell types), indicating that gene dysregulation associated with chronic HIV may occur across many immune cell types. Though informative, these data from DICE do not include HIV status information to directly assess cell-type specific differential gene expression for the bulk tissue DEGs. Single-cell RNA sequencing studies that compare people living with chronic HIV to people without HIV are needed to further dissect chronic HIV impact on gene expression across and within immune cell types. Similar work in acute HIV infection for PLWH not undergoing ART has provided insight regarding variability in immune response across cell types and individuals^77^.

### DEGs and genetic susceptibility

We considered whether our data supported gene-level findings from genomic studies of HIV acquisition since those studies also compare PLWH to people without HIV. Powell et al.^78^ and Duarte et al.^79^ conducted genetic association analyses that reported genes expressed in blood that were related to genetic risk of HIV acquisition (*EFCAB14*, *HERC1, HIST1H4L, IER3*). None were identified as DEGs in our study, suggesting that these potential HIV susceptibility genes are not persistently differentially expressed in PLWH on ART. Additionally, our partitioned heritability analysis did not show enrichment of genetic associations for HIV acquisition at genomic loci surrounding HIV-associated DEGs, and significant eQTLs for DEGs did not colocalize with genetic signal for HIV acquisition. A lack of heritability enrichment was also noted for all the cancer traits and most immune disorders we evaluated, which supports a hypothesis that the differential gene expression observed in this study is a result of living with chronic HIV infection while on ART and not a shared genetic predisposition with HIV acquisition or various cancers. In contrast, significant heritability enrichment at HIV-associated DEGs for Crohn’s Disease, asthma, rheumatoid arthritis, primary biliary cirrhosis, and primary sclerosing cholangitis implies that transcriptional alterations to these DEGs may impact pathogenesis and progression for these diseases. Prior studies have observed differences in incidence or activity of these diseases among PLWH and/or on ART compared to individuals not living with HIV^80–83^. For example, in Crohn’s disease, one hypothesis is that decreased CD4 T cell counts resulting from HIV infection attenuates the severity of inflammatory bowel disease^84,85^.

We identified colocalization of genetic signals among several traits and eQTLs for HIV-associated DEGs, despite the general lack of heritability enrichment for most of the traits we evaluated. The genes that exhibited eQTL colocalization with pancreatic cancer, *FAM135B* and *RP11-826N14.7*, are not well-characterized. The latter encodes a long noncoding RNA, which are increasingly studied for their role in cancer development^86^. *CMTM8*, whose eQTL colocalized with lymphocytic leukemia, has been noted in the regulation of apoptosis and cancer inhibition for other cancer types^87^. Several other HIV-associated DEGs had eQTLs colocalized with at least one immune-related trait. *ANKRD55*, whose eQTL colocalized with three autoimmune disorders assessed in this study (systemic lupus erythematosus, rheumatoid arthritis, and atopic dermatitis), has gene expression or variants within and proximal to the gene body previously linked to multiple autoimmune disorders, including rheumatoid arthritis and systemic lupus erythematosus^32^. Similar observations from past studies were made for *CCR6*^88^, *IKZF3*^89,90^, *ITGA4*^91,92^, and *LIME1*^93–95^—additional genes with colocalization signal with immune-related traits in this study. We only observed *SESN1* eQTLs colocalizing with primary sclerosing cholangitis and asthma, but this gene has been associated with carcinogenesis through its relationship as a p53-mediated mTOR pathway inhibitor^96,97^. Similarly, *FUT11* eQTL colocalization was only observed with type 2 diabetes and primary biliary cirrhosis, but this gene has been linked to regulation of the mTOR pathway, pancreatic cancer, and hepatocellular carcinoma^98,99^.

### DEGs as targets of pharmacological treatment

The identification of 111 HIV-associated DEGs as targets for clinical trial or approved drugs provides a survey of potential compounds that could alter the pathways overrepresented for DEGs, such as the cancer-related pathways. The example of midostaurin and bryostatin 1 as drugs that both target the ART-affected, HIV-associated, eGene *PRKCA* showcases the complex interactions to consider when assessing drug efficacy and safety in PLWH. One could hypothesize that midostaurin and bryostatin 1 have a genotype-dependent efficacy that would be further modulated if both drugs were simultaneously introduced within PLWH that are on ART. Ideally the interactions would be synergistic, but even so this would have implications for proper dosing. Furthermore, the antiviral properties of bryostatin 1 and its link to midostaurin through *PRKCA* as a common target suggests midostaurin could have an impact on HIV latency/latency reversal in PLWH. As more PLWH reach older age and polypharmacy becomes more common^100^, identifying and understanding genotype-dependent dosage effects, drug-drug interactions, and how drugs can alter disease risk for common co-occurring conditions becomes increasingly important. This complexity notwithstanding, our results provide a large number of drug – target pairs that bare further study as potential points of intervention to reduce the risk for development of non-AIDS defining cancers.

### Conclusion

In this first transcriptome-wide study of virally suppressed PLWH on ART, these findings highlight wide-spread gene dysregulation. Among these genes modulated by living with HIV and ART are numerous links to altered function in various cancers or immune-related diseases. These genes, and potentially drugs that target them, may be key to developing treatments for co-occurring health conditions in PLWH. Until a cure for HIV infection is developed, understanding the health implications of transcriptomic changes, genetics, drug-drug interactions, and ART while living with HIV will remain a critical component to ensuring PLWH receive appropriately tailored healthcare and to reducing co-occurring conditions associated with living with HIV.

## Online Methods

### Cohort description

Phenotypic and demographic data, RNA samples, and DNA samples were obtained from individuals of European ancestry originating from two cohorts with harmonized data for PLWH and people seronegative for HIV-1 who inject drugs in Vancouver, Canada (The AIDS Care Cohort to evaluate Exposure to Survival Services [ACCESS] and the Vancouver Injection Drug Users Study [VIDUS]). Details of cohort design and recruiting for the ACCESS and VIDUS cohorts have been described in detail elsewhere^101^. Phenotype, genotype, and gene expression data from 361 PLWH classified as virally non-detectable (<200 viral copies/ml of blood sample) and 194 individuals without HIV were used for this study. We refer to the combined studies as the Vancouver People Who Inject Drugs Study (VPWIDS). Biological samples obtained from VPWIDS are denoted as “HIV+ samples” when obtained from PLWH and “HIV-samples” when obtained from study participants living without HIV. Participants provided informed consent using methods approved by the Institutional Review Boards of University of British Columbia – Vancouver and RTI International.

### RNA sequencing and data processing

RNA was extracted from whole blood samples of VPWIDS participants and prepared for sequencing using the NuGEN Universal Plus mRNA-seq library preparation kit with human globin AnyDeplete primers. RNA samples were sequenced using paired-end 75 bp reads on an Illumina NextSeq by the Rutgers University Cell and DNA Repository (RUCDR Infinite Biologics). Following RNA sequencing (RNA-seq), read adapters were trimmed using cutadapt v1.15^102^, and reads less than 20 bp were removed using sickle v1.33 (https://github.com/najoshi/sickle). To generate quality control metrics, adapter trimmed RNA-seq reads were mapped to the GRCh38 human reference transcriptome (GENCODE v28) using HISAT2 v2.1.0^103^. Transcript-level quantifications were estimated using Salmon v0.11.2^104^. Gene-level quantifications were derived from the transcript-level estimates using the R package, *tximport*^105^.

To identify potential sample swaps, genotype correlations derived from genotyping array and RNA-seq data were calculated. Genotypes were called from RNA-seq alignments using the *mpileup* utility from *SAMTools*^106^. Problematic samples were identified by calculating pairwise genotype correlations (Pearson’s *r*) between the RNA-seq and genotype datasets. Samples were either excluded or mismatches resolved if r>0.8 for non-matching sample IDs or r<0.6 for matching sample IDs.

Sex discrepancies were assessed following the resolution of sample swaps. Males and females formed two distinct chromosome Y gene expression clusters, resulting in a bimodal distribution of normalized per-sample mean count values across chromosome Y genes. Consequently, discrepant sex assignments based on k-means (k=2) clustering of the mean count values of chromosome Y genes were excluded. The following sample-level exclusions were applied: low transcript diversity (unique transcripts detected per-sample) defined as the lower quartile - 1.5*IQR; read duplication rate >=0.75; missing genotype data; problematic based on sample swap analysis; sex discrepancies. RIN >5 and ribosomal RNA mapping rate were also considered as quality filters but did not result in any additional sample exclusions.

### Differential gene expression analysis

Differential gene expression by HIV status was tested using RNA-seq data from 361 PLWH and 194 people living without HIV. Prior to model fitting, genes considered lowly expressed were excluded (≤10 counts in ≤30% of samples [the approximate proportion of the smaller HIV status group]). Using the 20,211 remaining genes, surrogate variables (SVs) were estimated with *SVA v3.34.0*^107^ on variance-stabilized, median-of-ratios normalized counts^108^ to capture sources of technical and biological gene expression variance orthogonal to variation by HIV status.

A bootstrap sampling approach was taken to identify differentially expressed genes (DEGs) by HIV status, because no whole blood data set with a comparable sample size and experimental design exists to conduct a suitable replication analysis. For each bootstrap iteration, 80% of the HIV+ and HIV-samples were randomly selected (i.e., stratified sampling) and analyzed with *DESeq2 v1.26.0*^109^ using a negative binomial generalized linear model with HIV status and 30 SVs as explanatory variables.

Shrinkage was applied to fold change estimates using *apeglm*^110^. After 1000 bootstrap iterations, we used the p-values across bootstrap iterations for a given gene to determine statistical significance. We expect robust signals to have p-value distributions with values mostly or exclusively below a stringent threshold. Given a p-value significance threshold α=0.05, we applied Bonferroni correction to obtain an adjusted alpha, α_adjusted_=2.47e-06 (α/20211) and calculated a permutation p-value from how often a gene’s p-value is smaller than α_adjusted_. We classified a gene as a DEG if ≥95% of its bootstrap p-values are less than α_adjusted_ (permutation p-value ≤0.05).

### Gene set overrepresentation analysis

Gene set overrepresentation analysis was conducted using the R package *clusterProfiler* (v4.2.0)^111^ with two gene set databases: Gene Ontology (GO) biological process terms and the Kyoto Encyclopedia of Genes and Genomes (KEGG) canonical pathway terms. GENCODE v28 IDs for DEGs were converted to ENSEMBL IDs for compatibility with GO and KEGG database IDs used by *clusterProfiler*. The semantic similarity measure proposed by Wang *et al.*^112^ was used to calculate pairwise semantic distances between significant GO terms (FDR <0.05). The semantic relationships among GO terms were represented in a two-dimensional coordinate space using UMAP^113^ followed by clustering using HDBSCAN^114^. These discrete groupings do not modify the GO term results but organize terms to aid in biological interpretation and categorization.

### Bulk gene expression deconvolution and differential cell-type proportion testing

To estimate cell-type proportions, median-of-ratios normalized transcripts per million (TPM) values were obtained and genes considered lowly expressed were excluded (≤10 counts in ≤30% of samples [the approximate proportion of the smaller HIV status group]). Expression data for genes that constitute that CIBERSORTx LM22 cell-type reference panel were input to CIBERSORTx^115^ to obtain cell-type proportion estimates for 22 blood cell types. CIBERSORTx was run with the following parameter specifications: B-mode batch correction enabled, quantile normalization disabled, absolute run mode specified, and 1000 permutation iterations. Expression data for genes overlapping with the LM22 source gene expression profile matrix (11,724 of 11,845 genes) was provided for use in B-mode batch correction.

Prior to differential cell-type proportion testing, we identified sample outliers using a CIBERSORTx quality metric for cell-type proportion estimation. This metric equates to the correlation between the original bulk tissue sample gene expression and the reconstituted bulk gene expression from the estimated cell-type proportions. Any sample with a quality metric smaller than three standard deviations below the mean were considered outliers and excluded (seven sample exclusions). To identify cell-type proportion estimates that differed by HIV status, a linear regression model was fit for each cell type with the cell type proportions as the outcome variables. The explanatory variables for all models included HIV status, sex, ever vs. never sexually transmitted disease status, opioid use status (whether heroin was used within the last six months), age at blood draw, and five genotype PCs. Two-sided *t*-tests were conducted to identify significant, non-zero coefficient estimates for HIV status. Cell-type proportions were considered significantly different by HIV status for Bonferroni adjusted *p* <0.05.

### Genotyping and imputation

DNA was extracted from whole blood samples and genotyped on Illumina Infinium OmniExpress-24 BeadChip arrays by The Genomics Center at Rutgers University. Samples were removed during quality control using the following criteria: genotype missing rate >3%, sample duplication (identity-by-state >90%), first-degree relatedness (identity-by-descent >40%), discordance between self-reported sex and genotype-derived sex (*F*_st_ < 0.2 for chromosome X single nucleotide polymorphisms to confirm females and *F*_st_ > 0.8 to confirm males, where *F*_st_ denotes the coefficient of inbreeding), excessive homozygosity (*F*_st_ > 0.5 or *F*_st_ < −0.2), discordance between self-reported race and genotype-derived ancestry (AFR ≥25% and EAS ≥25% when using STRUCTURE^116^ with 1000 Genomes Phase 3 reference panel AFR, EUR, and EAS superpopulations^117^) or chromosomal anomalies. Genetic variants were removed due to missing rate >3% across samples or Hardy-Weinberg equilibrium *P* < 1 × 10^−4^. Following quality control, phasing and imputation were performed with the TOPMed Imputation Server^118^ using Eagle v2.4^119^, Minimac4^120^, and the TOPMed reference panel (genome build GRCh38)^121^. Imputed variants with R-squared <0.8 and minor allele frequency ≤1% were removed.

### Genotype PCA

Genotype principal components were derived using linkage disequilibrium (LD)-pruned observed genotypes (i.e., pre-imputation). Regions of high LD (GRCh37 coordinates chr5:43964243-51464243; chr6:24892021-33392022; chr8:7962590-11962591; chr11:45043424-57243424) were first removed, followed by LD-pruning using PLINK v1.9^122^ with the option “*--indep-pairwise 1500 150 0.2”*. The top 10 principal components were then computed using EIGENSTRAT v7.2.1^123^.

### Expression quantitative trait loci mapping

For HIV-associated DEGs, we tested whether imputed genetic variants in *cis* were expression quantitative trait loci (eQTL) using the following steps: 1) latent factor estimation and selection, 2) *cis* window size selection, 3) linear regression model fitting and association testing, and 4) multiple testing correction. Across steps, we used the following linear model structure with Matrix eQTL^124^ for variant-gene association testing:

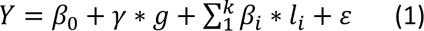

Here, *Y* is the median-of-ratios normalized counts for a given gene (additionally transformed using a rank-inverse normal transformation in step 3), *g* is the imputed genotype dosage, and *l_i_* are latent factors estimated by PEER^125^ to capture sources of technical and biological gene expression variance not attributable to HIV status or genetic factors. The *γ* and *β_i_* terms are the effect size estimates for the genotype dosage and latent factors, respectively, and *ε* is the error term. Matrix eQTL tests and reports for the significance of *γ*, with the null hypothesis *γ* = 0 and the alternative *γ* ≠ 0.

Latent factors were selected similarly to the “standard approach” as described by Fan *et al.*^126^ Briefly, a variance-stabilizing transformation was applied to *Y*. These transformed counts and an HIV status indicator variable were provided as inputs to PEER for estimating 50 latent factors, ordered by decreasing variance. For variants within a 2 megabase (Mb) total window (i.e., 1 Mb upstream and downstream) around chromosome 1 genes, eQTL were mapped for 50 linear models based on equation 1. Each model used a different number of latent factors, *k*, with *k* ∈ {1, 2, …, 50}. For each of the models, gene-level p-values were assigned by choosing the smallest p-value across association tests for a given gene. The number of significant eQTL genes (eGenes) from each model was determined through permutation testing. Genotype dosages were permuted 25 times for each gene for the 50 models to create null, gene-level p-value distributions. We classified eGenes as any gene with ≥95% permutation p-values larger than the non-permutation p-value. By assessing the number of eGenes discovered for each of the 50 models (i.e., eGenes discovered as a function of the numbers of latent factors), 13 factors were selected to balance model complexity with maximizing eGene discovery (Supplemental Figure 5A).

To select a *cis* window size for association testing with the final model, we first applied equation 1 with *k* = 13 for eQTL mapping of genetic variants within a 2 Mb total window around an HIV status DEG. We observed a systematic decrease in −log_10_ p-values with increasing variant distance from a gene (Supplemental Figure 5B). Using the approximate inflection point for −log_10_ p-values as a function of variant distance from a gene, we selected a final model total *cis* window size of 800 kb.

After *cis-*eQTL mapping using equation 1 with *k* = 13 and a window size of 800 kb, p-values were corrected for multiple hypothesis testing using a two-stage approach. This hierarchical approach first accounts for association tests across variants for a given gene, then corrects for tests across all DEGs. In the first stage, all nominal p-values for a given gene were adjusted using EigenMT^127^ with parameters *-- var_thresh 0.99*, *--window 200*, and *--cis_dist 400000*. In the second stage, EigenMT adjusted p-values were multiplied by the number of genes tested as a final correction. A two-stage adjusted p-value cutoff of 0.05 was used to identify statistically significant eQTL.

### Partitioned heritability analysis

We used stratified LD score regression (s-LDSC) v1.0.1^128^ with summary statistics from 30 genome-wide association studies (GWAS) (see Supplemental Table 7 for GWAS details) to test for trait heritability enrichment of genetic variants proximal (up to 100 kb total *cis* window size) to DEGs by HIV status. Aside from GRCh37 genomic coordinates for HIV status DEGs and the 50 kb up and downstream regions around them, several input files for s-LDSC are required that can be obtained from the developers at https://console.cloud.google.com/storage/browser/broad-alkesgroup-public-requester-pays/LDSCORE. Specifically, the *--bimfile* and --*bfile* parameters (data for variants used in LD score calculations) used files from *1000G_Phase3_plinkfiles.tgz*, *--ref-ld-chr* (LD scores) used files from *1000G_Phase3_baseline_ldscores.tgz*, *--w-ld-chr* (LD score regression weights) used files from *weights_hm3_no_hla.tgz*, and *–frqfile-chr* (allele frequencies) used files from *1000G_Phase3_frq.tgz*. These files contain data from the 1000 Genomes project Phase 3 EUR superpopulation.

### Colocalization of GWAS signals and HIV eQTL

For each HIV-associated DEG with a significant eQTL, colocalization was tested using the *coloc v5.1.0* R package^129^ with eQTL summary statistics for variants proximal (800 kb total *cis* window size) to the DEG and summary statistics from the same 30 GWAS used in partitioned heritability analysis. Each DEG in the eQTL data was specified as a quantitative trait for the *coloc.abf* function, with the sample standard deviation parameter calculated from the RINT and median-of-ratios normalized gene counts. GWAS traits were specified for *coloc.abf* as case-control if the outcome variable was categorical and binary and quantitative otherwise. Proximal variants were subset to only those present in both the eQTL and GWAS data and minor allele frequencies for these variants were calculated based on the imputed genotype dosages for the study cohort. A posterior probability of colocalization (*coloc* hypothesis H4) >0.75 was used to denote a colocalization event between a given eQTL and GWAS locus.

## Supplemental Table Legends

**Supplemental Table 1.** Summary statistics for differential gene expression analysis by HIV status. Bootstrapping was applied to fit 1000 differential expression regression models, resulting in 1000 sets of summary statistics that are summarized in this table as median/median absolute deviation and mean/standard deviation values. The ART affection status is only provided for significant DEGs. “NA” values indicate missing values.

**Supplemental Table 2.** GO Biological Process terms with significant overrepresentation of HIV status DEGs (FDR <0.05) determined using the R package *clusterProfiler*. The gene ratio denotes the number of HIV status DEGs overlapping a given GO term divided by the number of DEGs present across all GO terms. The background ratio indicates the number of genes linked to a given GO term divided by the total number of genes represented across all GO terms. The *Overlapping DEGs* values include ENSEMBL IDs delimited by “/” for DEGs overlapping a given GO term.

**Supplemental Table 3.** MSigDB KEGG canonical pathway terms with significant overrepresentation of HIV status DEGs (FDR <0.05) determined using the R package *clusterProfiler*. The gene ratio denotes the number of HIV status DEGs overlapping a given KEGG term divided by the number of DEGs present across all KEGG terms. The background ratio indicates the number of genes linked to a given KEGG term divided by the total number of genes represented across all KEGG terms. The *Overlapping DEGs* values include ENSEMBL IDs delimited by “/” for DEGs overlapping a given KEGG term.

**Supplemental Table 4.** GO Biological Process and MSigDB KEGG canonical pathway terms with significant overrepresentation of ART-affected, HIV status DEGs (FDR <0.05) determined using the R package *clusterProfiler*. The gene ratio denotes the number of HIV status DEGs overlapping a given GO/KEGG term divided by the number of DEGs present across all GO/KEGG terms. The background ratio indicates the number of genes linked to a given GO/KEGG term divided by the total number of genes represented across all GO/KEGG terms. The *Overlapping DEGs* values include ENSEMBL IDs delimited by “/” for DEGs overlapping a given GO/KEGG term.

**Supplemental Table 5.** GO Biological Process and MSigDB KEGG canonical pathway terms with significant overrepresentation of ART-unaffected, HIV status DEGs (FDR <0.05) determined using the R package *clusterProfiler*. The gene ratio denotes the number of HIV status DEGs overlapping a given GO/KEGG term divided by the number of DEGs present across all GO/KEGG terms. The background ratio indicates the number of genes linked to a given GO/KEGG term divided by the total number of genes represented across all GO/KEGG terms. The *Overlapping DEGs* values include ENSEMBL IDs delimited by “/” for DEGs overlapping a given GO/KEGG term.

**Supplemental Table 6.** ART-unaffected, HIV status DEGs that overlap with GO Biological Process and MSigDB KEGG canonical pathway terms that had significant overrepresentation of ART-affected DEGs (FDR <0.05).

**Supplemental Table 7.** GWAS traits included in partitioned heritability enrichment and colocalization analysis.

**Supplemental Table 8.** Partitioned heritability enrichment analysis summary statistics output from s-LDSC. Results are shown for testing heritability enrichment of 30 GWAS traits at regions within and 100 kb proximal to HIV status DEGs.

**Supplemental Table 9.** Summary statistics for significant variant-gene associations (adjusted p-value <0.05) from eQTL mapping for HIV status DEGs. Indicator variables are included for a DEG to denote ART affection status and overlap with the MSigDB KEGG canonical pathways term *pathways in cancer*.

**Supplemental Table 10.** Colocalization analysis results for HIV status DEGs and 30 GWAS traits. Each value in the table corresponds to the *coloc* hypothesis 4 (H4) posterior probability for a given DEG and GWAS trait. The H4 posterior probability is defined as the probability that both traits (i.e., the GWAS trait and gene expression for the DEG) are associated and share a single causal variant.

**Supplemental Table 11.** Drugs from DrugBank, PHAROS, Open Targets, or TTD that target an HIV status DEG. The drug ID corresponds to the ID used by the corresponding drug database for a given record.

Some drugs have only an ID and no name. These are denoted with the value “NA” for the drug name. Indicator variables are included for a DEG to denote ART affection status, overlap with the MSigDB KEGG canonical pathways term *pathways in cancer*, and whether the DEG has a significant eQTL.

## Author Contributions

Author contributions are categorized using the CRediT taxonomy (credit.niso.org).

BCQ: Formal analysis, methodology, software, visualization, writing – original draft, writing – review & editing

EE: Formal analysis, methodology, software, writing – review & editing

LZ: Formal analysis, software, writing – review & editing

CW: Formal analysis, software, writing – review & editing

JAM: Formal analysis, software, writing – review & editing

JKS: Formal analysis, writing – review & editing

FF: Writing – review & editing

LJB: Writing – review & editing

MJSM: Resources, writing – review & editing

KH: Resources, writing – review & editing

KD: Resources, writing – review & editing

DBH: Conceptualization, writing – review & editing

KX: Conceptualization, funding acquisition, writing – review & editing

BEA: Conceptualization, funding acquisition, writing – review & editing

EOJ: Conceptualization, funding acquisition, project administration, supervision, writing – review & editing

## Data Availability

All data produced in the present study are available upon reasonable request to the authors.

## Acknowledgements

This work was funded by the National Institutes of Health grants U01DA038886, U01DA021525, R61/R33DA047011, R01DA051908, and R01DA051908-03S1.

